# Large-scale cerebrospinal fluid proteomic analysis in Alzheimer’s disease patients reveals five molecular subtypes with distinct genetic risk profiles

**DOI:** 10.1101/2023.05.10.23289793

**Authors:** Betty M Tijms, Ellen M Vromen, Olav Mjaavatten, Henne Holstege, Lianne M Reus, Sven van der Lee, Kirsten EJ Wesenhagen, Luigi Lorenzini, Lisa Vermunt, Vikram Venkatraghavan, Niccoló Tesi, Jori Tomassen, Anouk den Braber, Julie Goossens, Eugeen Vanmechelen, Frederik Barkhof, Yolande AL Pijnenburg, Wiesje M van der Flier, Charlotte E Teunissen, Frode Berven, Pieter Jelle Visser

**Affiliations:** Alzheimer Center Amsterdam, Neurology, Vrije Universiteit Amsterdam, Amsterdam UMC location VUmc, Amsterdam, the Netherlands; Amsterdam Neuroscience, Neurodegeneration, Amsterdam, the Netherlands; PROBE, department of biomedicine, University of Bergen, Bergen, Norway; Department of Clinical Genetics, Vrije Universiteit Amsterdam, Amsterdam UMC location VUmc, Amsterdam, the Netherlands; Center for Neurobehavioral Genetics, Semel Institute for Neuroscience and Human Behavior, David Geffen School of Medicine, University of California Los Angeles, Los Angeles, USA; Genomics of Neurodegenerative Diseases and Aging, Human Genetics, Vrije Universiteit Amsterdam, Amsterdam UMC location VUmc, Amsterdam, the Netherlands; Department of Radiology and Nuclear Medicine, Amsterdam UMC location VUmc, Vrije Universiteit Amsterdam, Amsterdam, the Netherlands; Amsterdam Neuroscience, Neuroimaging, Amsterdam, the Netherlands; Neurochemistry Laboratory, Department of Clinical Chemistry, Amsterdam UMC location VUmc, Vrije Universiteit Amsterdam, Amsterdam, the Netherlands; Delft Bioinformatics Lab, Delft University of Technology, Delft, The Netherlands; ADx NeuroSciences, Ghent, Belgium; Institutes of Neurology and Healthcare Engineering, University College London, London, UK; Alzheimer Center Limburg, School for Mental Health and Neuroscience, Maastricht University, Maastricht, the Netherlands; Department of Neurobiology, Care Sciences and Society, Division of Neurogeriatrics, Karolinska Institutet, Stockholm, Sweden

**Keywords:** Alzheimer’s disease, cerebrospinal fluid, proteomics, disease heterogeneity

## Abstract

Alzheimer’s disease (AD) is heterogenous on the molecular level. Understanding this heterogeneity is critical for AD drug development. We aimed to define AD molecular subtypes by mass spectrometry proteomics in cerebrospinal fluid (CSF). Of the 3863 proteins detected in CSF, 1058 proteins had different levels in individuals with AD (n=419) compared with controls (n=187). Cluster analyses of AD individuals on these 1058 proteins revealed five subtypes: subtype 1 was characterized by neuronal hyperplasticity; subtype 2 by innate immune activation; subtype 3 by RNA dysregulation; subtype 4 by choroid plexus dysfunction; and subtype 5 by blood-brain barrier dysfunction. Distinct genetic profiles were associated with subtypes, e.g., subtype 1 was enriched with *TREM2 R47H*. Subtypes also differed in brain atrophy and clinical outcomes. For example, survival was shorter in subtype 3 compared to subtype 1 (5.6 versus 8.9 years). These novel insights into AD molecular heterogeneity highlight the need for personalized medicine.

---

Alzheimer’s disease (AD) is the leading cause of dementia, affecting about 44 million people worldwide^1^. AD is histopathologically defined by amyloid plaques and hyperphosphorylated tau tangles in the brain^2^, but its underlying pathophysiology remains largely unclear. Genetic studies, as well as brain tissue gene expression and proteomic studies have indicated that many different pathophysiological processes are associated with amyloid and tau pathology, including but not limited to synaptic plasticity, the innate immune system, neuroinflammation, lipid metabolism, RNA metabolism, the matrisome, and vascular function^3–9^. This heterogeneity may explain why previous AD trials had no or limited clinical effects^10–12^. For example, we previously found that CSF BACE1 levels were abnormally increased in a specific AD subtype, suggesting that BACE inhibition may be effective in a subgroup only^9, 13^. This highlights the need for personalized treatments and for in-vivo tools to define such molecular subtypes.

CSF is the most accessible biofluid to study molecular complexity of neurodegenerative diseases during life: CSF is in close contact with the brain, and protein concentrations in CSF reflect the brain’s ongoing (patho)physiological processes. We previously discovered and replicated three distinct molecular AD subtypes through investigation of respectively 707 and 204 proteins in CSF^9^. The proteins involved in these subtypes represent distinct biological processes such as neuronal plasticity, innate immune activation and blood-brain barrier dysfunction^9^. Subtype specific molecular alterations were already present at a very early stage of AD, when cognition was still intact and neuronal damage still limited. Furthermore, these molecular processes were previously also identified in AD post-mortem tissue proteomic studies ^3, 4, 6, 8^. This supports the value of CSF proteomics to detect AD pathophysiological processes in living patients^4^.

Proteomic techniques have greatly improved since then, and can now detect thousands of proteins in CSF, providing an unprecedented opportunity to dissect the molecular processes associated with AD in detail. In our current study, we took advantage of these novel techniques in a new cohort and detected more than 3000 proteins in CSF. We also increased the number of individuals to 609 individuals to replicate and refine existing subtypes, to test if the higher complexity allows us to uncover more AD subtypes, and to study underlying genetic factors of these subtypes.

In our previous studies, we compared CSF AD subtypes on *APOE* e4 carriership (the strongest genetic risk factor for sporadic AD)^9, 14^ and on AD polygenic risk scores. In the current study we further extent genetic analyses, and compared subtypes on AD risk variants from a recent GWAS^5^. Moreover, we enriched for the relatively rare *TREM2* R47H and R62H mutations, as these are associated with a resp. 2.3 and 1.4-fold increased risk of AD^5^. *TREM2* R47H and R62H are supposed to impair microglia activation in AD^15^. We therefore hypothesized that carriers of *TREM2* variants could group together in a subtype with impaired microglial activation. A small number of patients (n=6) were carrier of autosomal dominant mutation in PSEN1 or APP and we performed exploratory analysis to which subtypes these genetic variants were associated.

To further characterize the subtypes, we compared the groups on clinical measures, atrophy patterns and enrichment for biological processes, transcription factors, and cell type specificity. This new large scale CSF proteomic study revealed five molecular AD subtypes. Three subtypes recapitulated our previously identified three subtypes (hyperplasticity, innate immune activation and blood-brain barrier dysfunction)^9^. We also identified two new AD subtypes: one with RNA dysregulation, and one with choroid plexus dysfunction. These subtypes were associated with distinct genetic risk profiles, further validating the biological underpinning of AD subtypes. The proteomic signatures associated with AD subtypes were present already in the preclinical stage and largely remained stable with increasing disease severity. Subtypes differed in the amount and pattern of cortical atrophy, cell type specific expression of proteins, vascular damage, progression rate from mild cognitive impairment to dementia and survival times in dementia.

Our results highlight the importance of neuronal plasticity, microglial impairment, innate immune activation, RNA processing choroid plexus and blood-brain barrier dysfunction in AD pathogenesis, and provide a comprehensive resource that informs on which proteins and pathways are dysregulated in particular subtypes of AD patients.

## Results

We analysed CSF samples from 609 individuals that were selected from Alzheimer Center Amsterdam related studies^16–19^ (see online methods). Of this sample, 419 had AD as defined by abnormal CSF amyloid beta 1-42 (abeta42) levels across clinical stages (i.e., 107 with normal cognition, 103 with mild cognitive impairment (MCI), and 209 with dementia). The 187 controls were required to have normal cognition and normal CSF abeta42 and tau levels. CSF proteins from each sample were enzymatically digested and the peptides were labelled with tandem mass tags (TMT), fractionated, and analysed by LC-MS/MS (see online methods). A total of 3863 proteins was identified, of which 1309 were observed across all individuals. We then selected proteins that different between controls and AD (S-table 2). We repeated those analyses stratified on tau-levels or disease stage, because protein levels can change in a non-linear way with these variables^13, 20^. This led to a total of 1058 AD-related proteins that were selected for cluster analyses (S-table 2). We then clustered AD individuals on AD-related proteins using the dual clustering approach ‘non-negative matrix factorisation’^21^ (figure 1). A particular strength of the algorithm is that individuals will per definition be allocated to one subtype, which is useful for diagnosis or patient stratification for trials.

**Figure 1.**
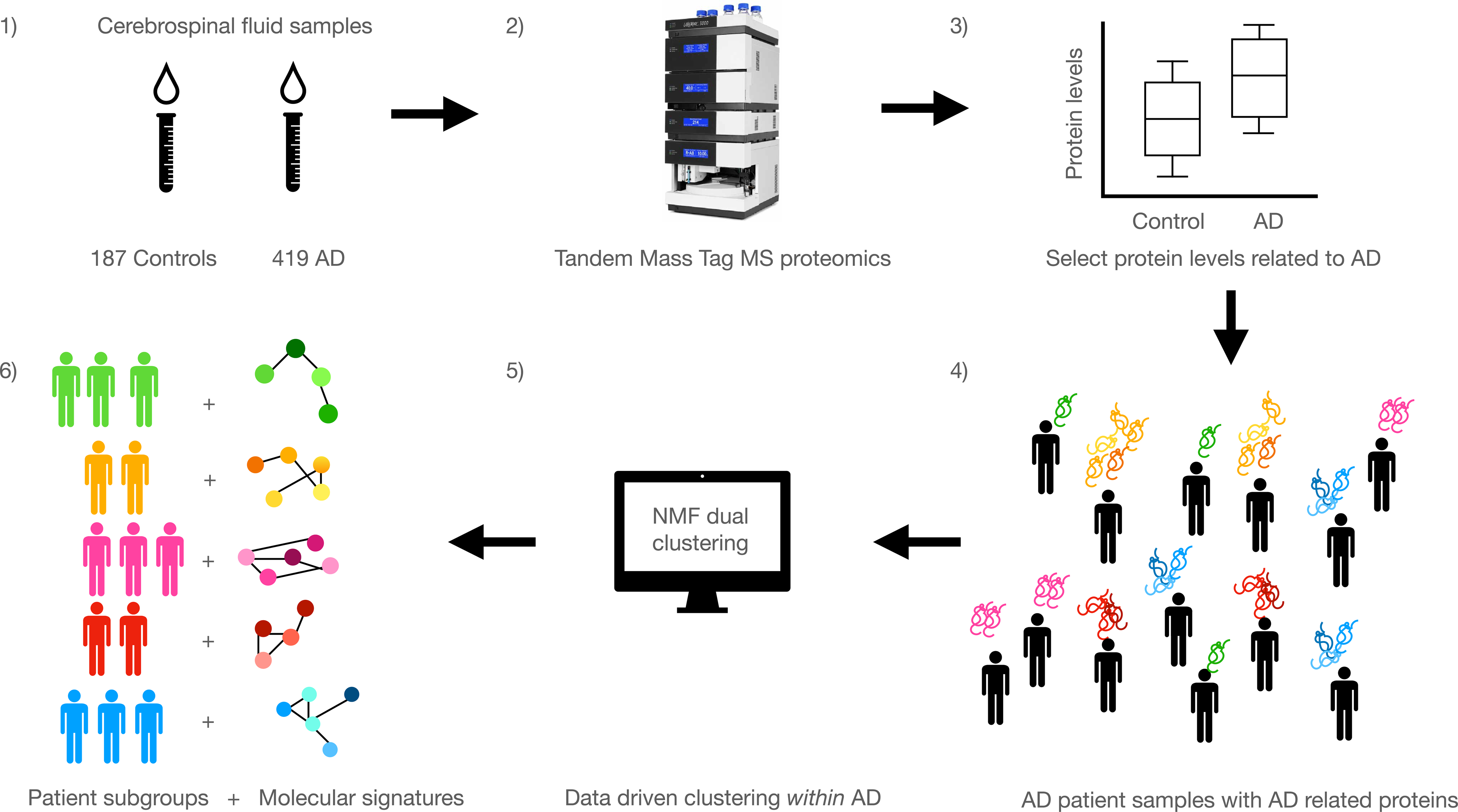
Schematic overview of subtype discovery within AD patients. 1) Cerebrospinal fluid (CSF) samples from 198 controls and 419 individuals were selected and 2) analysed with Alzheimer’s disease (AD) were analysed with tandem mass tag mass spectrometry to obtain untargeted proteomics. 3) Protein levels were then compared between controls and AD to select proteins associated with AD. 4) Within the AD group for proteins related to AD, data driven clustering was performed with non-negative matrix factorisation (NMF) (5). 6) The resulting patient subgroups were then molecularly characterised based on their corresponding proteomic signatures, and compared on clinical and biological characteristics.

### Five AD subtypes with distinct differences in clinical, molecular and genetic characteristics

Patients’ proteomic profiles clustered into 5 subtypes (figure 2a; S-table 3 for fit statistics): Subtypes 1, 2, and 5 recapitulated our previously detected subtypes with neuronal hyperplasticity (subtype 1), innate immune activation (subtype 2) and blood-brain barrier dysfunction (subtype 5). Additionally, two new subtypes emerged: one with RNA dysregulation (subtype 3) and one with choroid plexus dysfunction (subtype 4). The next sections discuss each subtype in detail on molecular, and genetic and clinical characteristics, here we briefly summarise these characteristics. Compared to controls, subtypes 1, 2 and 3 had increased CSF t-and p-tau levels, while subtype 4 and 5 had mostly normal tau levels (table 1). Subtypes differed in clinical stage, sex, and age, and so all subsequent analyses took these characteristics into account. Compared to controls, subtypes differed in rates of progression from MCI to dementia, with subtypes 2 and 5 having the highest risk, and subtype 4 the lowest (figure 3d), albeit not significantly different between subtypes (S-table 10a). Subtype 3 individuals with dementia had the most aggressive disease course of 5.6 years, which was shorter than subtype 1 with the longest survival time of 8.9 years (p=0.04; S-table 10b). These results suggests that different underlying molecular aspects may explain variability in decline. Analysis of MRI scans in individuals with dementia (n=159) indicated that subtypes differed in the degree and anatomical location of cortical atrophy (figure 2c; S-table 9). All subtypes had a higher prevalence of the APOE e4 genotype than controls, and a higher AD polygenic risk score, supporting their underlying AD genetic risk architecture. Subtypes had, however, different AD genetic risk profiles.

**Figure 2.**
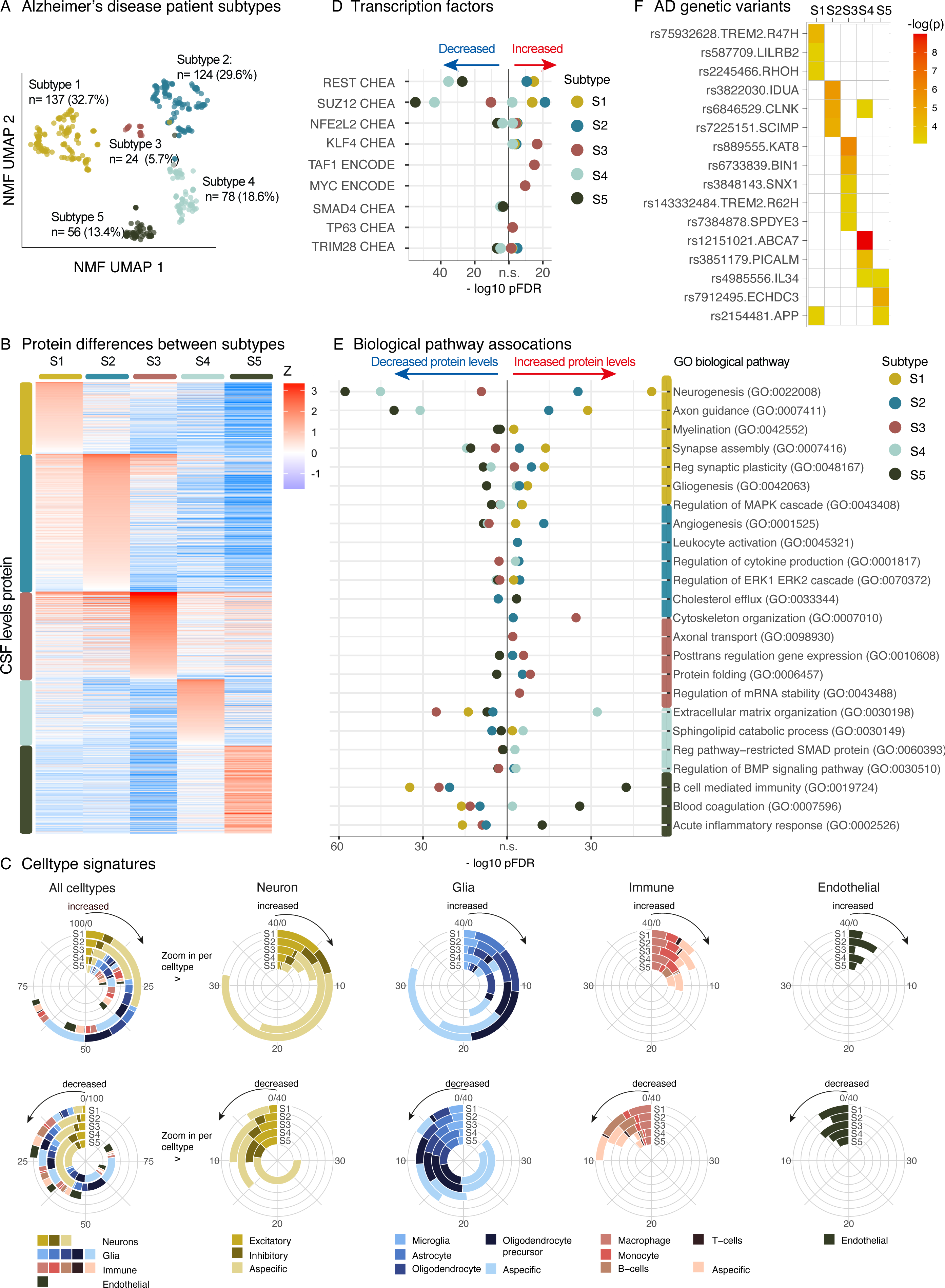
**A** Patient subtypes projected to UMAP space. **B** CSF protein levels averaged across individuals within subtypes. **C** Cell type specificity signatures for proteins associated with AD subtypes for proteins with increased levels on the top row, and decreased levels in the bottom row. Most left circle diagram shows all cell types associated with a subtype combined (most left circle diagrams). Proteins that could not be assigned to a specific cell type were not plotted (missing bar to 100% in first column). Circle diagrams to the right zoom into subcategories of specific celltypes (neurons, glia, immune cells and endothelial cells). Cell type specificity was determined according to the Human Protein Atlas. **D** Top transcription factors associated with subtypes from the CHEA and ENCODE databases. **E** GO biological pathways associated with subtypes (see supplemental material for all pathways). **F** AD genetic risk factors associated with specific subtypes, white indicates not significant. In all figures S1 is subtype 1 (hyperplasticity), S2 is subtype 2 (innate immune activation), S3 is subtype 3 (RNA dysregulation), S4 is subtype 4 (choroid plexus dysfunction), and S5 is subtype 5 (blood-brain barrier dysfunction). Supplementary tables 5, 6, 7, 8a and 8b list all proteins, pathways, transcription, and genetic factors tested with their statistical metrics.

**Figure 3.**
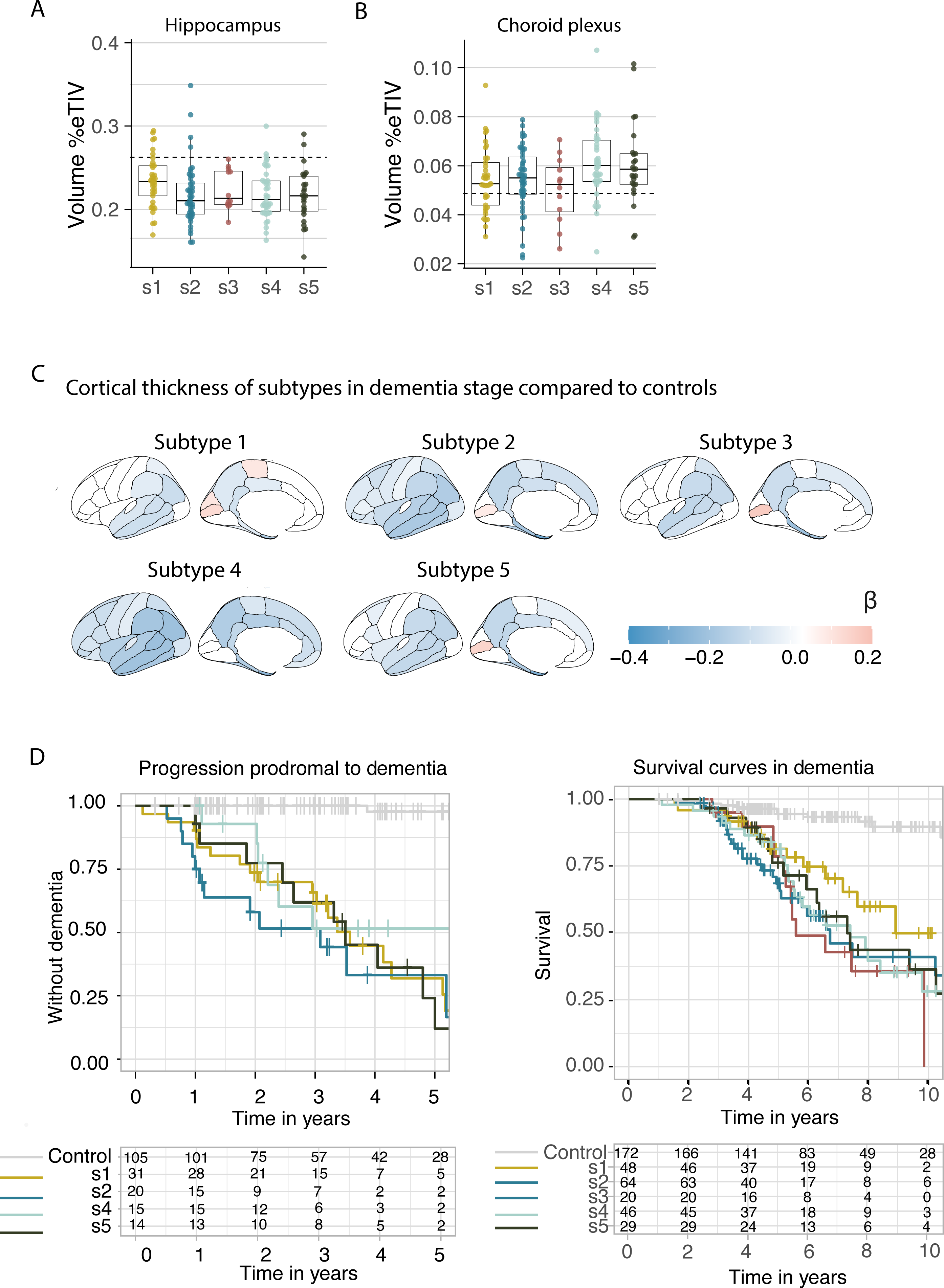
**A** Hippocampal volume compared between subtypes. **B** Choroid plexus volume compared between subtypes. **C** Cortical atrophy associated with AD subtypes as compared to controls. **D** Clinical progression from MCI to dementia according to subtype (left; excluding subtype 3 due to n=2), and time from dementia to death according to subtypes (right). All atrophy measures are based on individuals with dementia only. See supplemental tables 9, 10a and 10b.

**Table 1.**
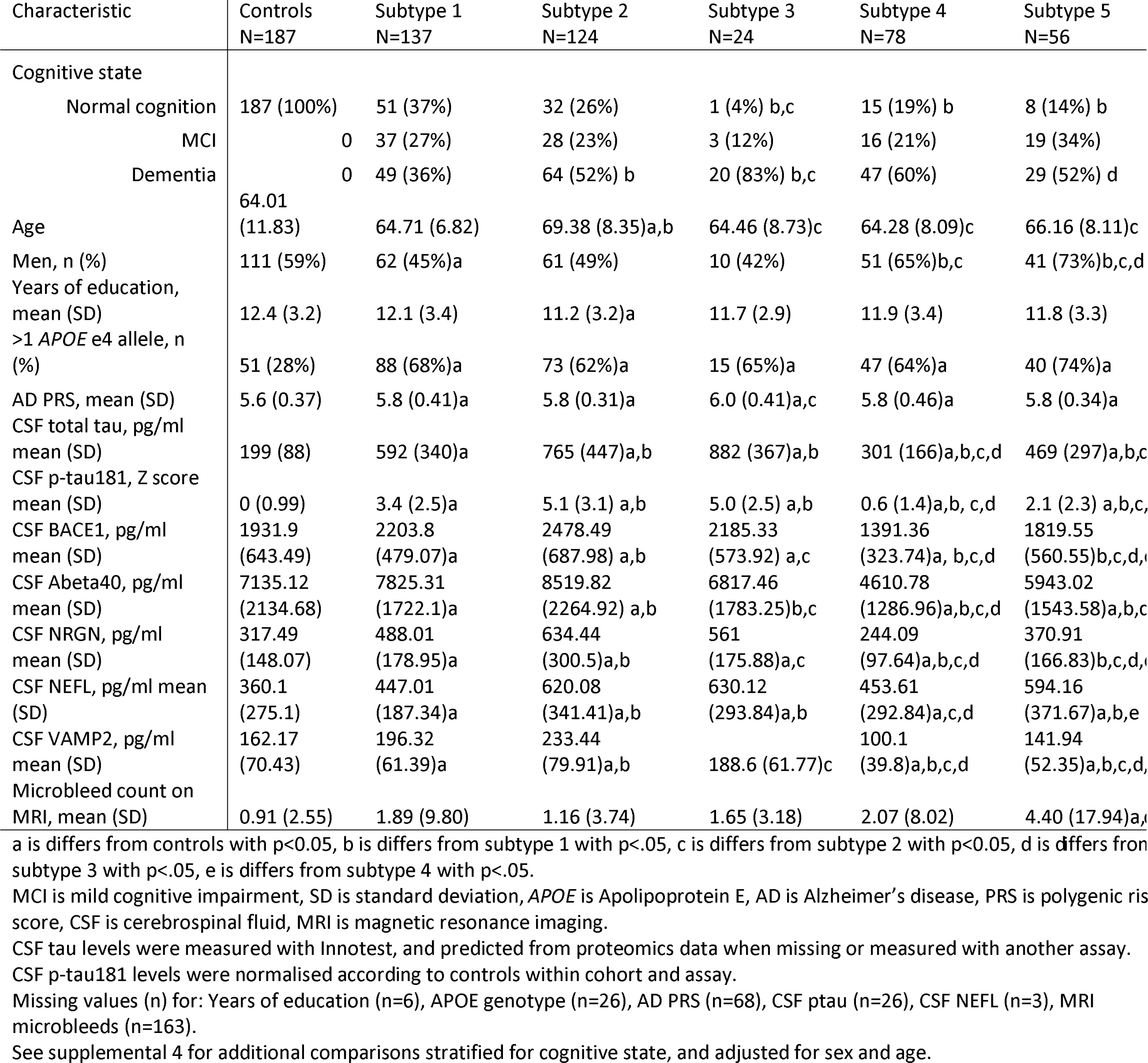
Comparison of subtypes on clinical characteristics

We next examined the molecular processes associated with AD subtypes. For each subtype we compared the levels of 2878 proteins against the control group (figure 2b; S-table 5). Proteins with different levels between a subtype and the control group were included in enrichment analyses to study associated biological processes and transcription factors. In order to aid comparability with e.g., gene expression literature we report gene names for proteins (UniProt codes are listed in S-table5). Stratification by disease stage resulted in similar differences to controls (correlations between 0.85-0.98; S-figure 1; S-table5 columns CY-HU), providing further support that AD subtypes reflect specific disease traits^9, 14^.

Below, we discuss the most distinct biological processes types, cell types AD risk variants and atrophy patterns that were associated with each subtype. Detailed results are reported in the supplemental material.

### Subtype 1: Hyperplasticity

Subtype 1 individuals (n=137, 32.7%) had compared to controls 827 proteins with increased CSF levels and 408 proteins with decreased levels. Of all subtypes, subtype 1 had the highest proportion of proteins specific for neurons, astrocytes, oligodendrocytes and oligodendrocyte precursor cells (figure 2c). Proteins with increased levels were associated with neuronal plasticity processes, including synapse assembly, axon guidance, neurogenesis, and gliogenesis (figure 2e, S-table 6). In addition, this neuronal hyperplasticity subtype had high BACE1 and amyloid beta 1-40 (abeta40) CSF levels, as well as high tau levels (table 1), like in our previous study^9^. While high tau levels were previously thought to reflect neuronal loss due to tangle formation, now more studies indicate that this may also reflect other processes^22, 23^. For example, hyperactive neurons and astrocytes have been reported to surround plaques^24, 25^. Neurons with increased activity secrete more amyloid as well as tau^26–30^, and fragments of those proteins may in turn drive hyperplasticity through enhanced gene transcription^31^. Indeed, proteins increased in subtype 1 included were enriched for the transcription factors REST (p_adjusted_=.018×10^-13^, figure 1d, S-table7) and SUZ12 (p_adjusted_ =.016×10^-12^), which regulate plasticity related processes through repression of neuronal differentiation genes^32, 33^. Previous studies pointed towards REST de-repression and increases of tau and plasticity related processes in AD brain tissue^8, 34^, iPSC neurons^35, 36^ and tau tangle carrying neurons^37^. Comparing subtype 1 increased proteins with those studies, we found an overlap 5 of 6 from^8^, 65 of 173 from^35^ and 46 of 127 from^37^ (S-table 5). Moreover, with the higher number of proteins that we measured compared to our previous study, we found other mechanisms that may contribute to the plasticity response observed in this subtype. For example, the lysosomal protein PLD3 was highest in subtype 1. High PLD3 levels have been reported in dystrophic neurites associated with ‘amyloid axonal spheroids’ ^38^. Such spheroids have previously been found to trigger axonal remodelling and local hyperactivity^38^. Furthermore, dystrophic neurites accumulate BACE1, which has been found to increase processing of APP^39^, and may explain the elevated BACE1 and abeta levels in this subtype.

Next, we tested which AD risk variants^5^ were more common in this subtype compared to controls. We found enrichment in subtype 1 for variants in *TREM2 R47H*, *LILRB2, RHOH, NCK2* and in *APP* (figure 2f; S-tables 8a, b). This subtype also included 3 of the 4 *PSEN1* carriers (S-table 8b). TREM2 is a transmembrane protein, of which the extracellular part binds ligands ^15^, including amyloid fibrils, that can activate microglia. The genetic variant *TREM2 R47H* has been associated with dampened microglia activation due to decreased ligand binding to TREM2’s extracellular part^40, 41^. *LILRB2* has been associated with a similar dampened immune activation as *TREM2*^42, 43^. Furthermore, *RHOH* and *NCK2* are signalling molecules downstream from TREM2 that influence cytoskeleton rearrangement of microglia, necessary to enable migration towards pathogens and amyloid plaques^44^. Normally, activated microglia form a tight barrier around plaques that decreases plaque surface, thereby minimizing plaque contact with neurites^15, 40, 45^. When microglial activation is dampened, such as observed in carriers of *TREM2* variants, amyloid plaques are less compact, with toxic oligomers sticking out that could damage nearby neurites^46–48^ and may lead to axonal dystrophy^47^, which may trigger a plasticity response. *TREM2* has also been implicated in impaired microglial synaptic pruning, which could further contribute to the hyperplasticity signature observed in this subtype^49–53^. Such an excess of synapses was previously associated with milder atrophy on MRI in *TREM2* mouse models^49^.

After analysing MRI in our data, we found that this subtype had the lowest degree of atrophy compared to the other subtypes (figure 3c; S-table9), and was restricted to the temporal and parietal lobes.

Together, our results provide further support for a hyperplasticity subtype in AD, which we now observe could be related to a dampened microglial response. Currently, therapies that boost TREM2 activation are in development^54^. We argue that individuals with this subtype may also respond to such treatments, even without carrying the *TREM2 R47H variant*.

### Subtype 2: Innate immune activation

Subtype 2 individuals (n=124, 29.6%) had compared to controls 986 proteins with increased CSF levels and 506 with decreased levels. A high proportion of proteins increased in subtype 2 was specific to microglia. Proteins with increased levels were associated with innate immune activation, including regulation of cytokine production. These included proteins from the complement complex (C1QA, C1QB, C1QC, C1S and C1R), as well as APOE and LPL. This subtype also had increased levels of microglial TAM receptors AXL and MERTK, and GAS6 (a MERTK ligand), which can detect and engulf plaques^55, 56^. We further found, for the first time, increased PYCARD levels specifically in subtype 2. PYCARD is also known as Apoptosis-associated speck-like protein containing a CARD (ASC), and is released by microglia with NLRP3 inflammasome activation^57, 58^. PYCARD can form ASC specks^59^, which are fibrils that worsen amyloid aggregation^58^ and induce tau phosphorylation^60^, providing a potential mechanism through which microglial activation may aggravate AD pathology. Indeed, subtype 2 individuals had higher p-tau levels than seen in subtype 1 (table 1). Other subtype 2 increased proteins were related to neuron-microglia signalling, such as CSF1, CSF1R, and CX3CL1.

Neuro-immune signalling occurs during normal neuronal development, during which microglia prune immature synapses^61–65^. However, activated microglia near diffuse and neuritic plaques may lead to excessive synaptic pruning^62^. This could lead to exacerbated atrophy on MRI as shown in mouse models^66, 67^. In line with those models, subtype 2 was one of the two subtypes with the most severe and widespread cortical atrophy on MRI compared to subtype 1, 3 and 5 (figure 3c; S-figure 2). Still, despite this severe atrophy, the levels of proteins related to neuroplasticity were increased in this subtype and these proteins overlapped with subtype 1, and were also enriched for the transcription factors REST and SUZ12. Possibly, the increase of plasticity related proteins may reflect an attempt to repair synaptic contacts, which succumbs in the presence of activated microglia. Alternatively, increased protein levels may reflect neuronal loss as a result of atrophy.

AD genetic variants associated with this subtype were *IDUA, CLNK*, and *SCIMP*, which are all implied in immune processes^5, 68, 69^.

Together, these results give detailed insight into the innate immune activation AD subtype, and suggest that an overactive innate immune system worsens the disease.

### Subtype 3: RNA dysregulation

Subtype 3 (n=24, 5.7%) emerged as a new subtype. Compared to controls this subtype had increased CSF levels for 516 proteins and decreased levels for 757 proteins. Proteins with increased levels were associated with cytoskeleton organisation, axonal transport, proteasome and protein folding (figure 2e; S-tables 5, 6). This subtype had the highest t-tau and NEFL CSF levels. BACE1 levels were higher than controls (table 1), but unlike subtypes 1 and 2, abeta40 levels were similar to controls, suggesting a different mechanism associated with higher BACE1 levels for this subtype. Proteins specifically increased in subtype 3 included heterogenous nuclear ribonucleoproteins (hnRNPs)^70^ and other RNA binding proteins, which may point to RNA dysregulation. HNRNPs, are involved in maturation of pre-mRNAs, mRNA stabilisation during transport and local mRNA translation for many RNAs, including those important for cytoskeleton organisation ^71, 72^. Disruptions in HnRNPs and mRNA have been associated with tau tangles in previous proteomic studies^73^. Mislocalised hnRNPs could lead to mis-and/or cryptic splicing, resulting in dysfunctional proteins^70, 74^. For example, cryptic splicing of STMN2 is a hallmark of TDP43 mislocalisation^75^, resulting in shorter proteins and decreased STMN2 levels in tissue^76^. In our data STMN2 was detected in a subset (n=84), and subtype 3 specifically had decreased levels of STMN2 compared to controls. Transcription factors associated with subtype 3 increased proteins were KLF4 (p_adjusted_=0.02×10^-15^), which is associated with axon regeneration ability^77^, as well as on TAF1 (p_adjusted_ =0.008×10^-13^) and MYC (p_adjusted_=0.02×10^-10^), which are interacting factors in cell differentiation processes^78, 79^. A previous study based on brain tissue gene expression found a similar AD subtype with increased TAF1 and MYC signalling and decreased synapse organisation^8^.

When testing AD risk factors, we found that subtype 3 was enriched for *BIN1*, which is known as ‘myc-box dependent interaction protein’. One of BIN1’s functions is to physically inhibit MYC^80^. BIN1 mainly localises in axons and has many isoforms arising from splicing^80^. BIN1 mis-splicing has been associated with de-inhibition of MYC and cytoskeleton disruptions^81^. *TREM2* R62H was also associated with this subtype. On MRI this subtype showed relatively mild atrophy like subtype 1, but also encompassed atrophy in the superior frontal gyrus. Other genetic risk variants associated with subtype 3 included *SPDYE3*, involved in the cell-cycle, *SNX1*, important for endosome sorting, and *KAT8* a lysine acetyltransferase^5, 69^.

While RNA dysfunction has been mainly observed in frontotemporal dementia^82^, we now find that these disruptions are associated with a specific AD subtype as well.

### Subtype 4: Choroid plexus dysfunction

Subtype 4 (n=78, 18.6%) was another new subtype. Compared to controls, this subtype had increased CSF levels of 467 proteins, and decreased levels of 626 proteins. A high proportion of proteins increased in subtype 4 were specific to microglia and other immune cells. Moreover, a large subset of proteins with increased levels (45%) was associated with high expression in the lateral ventricle choroid plexus (S-table 5), including TTR, SPARC, and extracellular matrix proteins such as DCN, LUM and COLA12. Biological processes associated with subtype 4 included cell adhesion, BMP and SMAD pathways, which are involved in choroid plexus development^83^ (figure 2e). The choroid plexus is located along the ventricles, where it produces CSF and is responsible for nutrient, lipid, and protein transfer across the blood-CSF interface^83^. It consists of a highly developed extracellular matrix that connects a dense vasculature to its epithelial cells^84^. On MRI, subtype 4 had the largest choroid plexus volume (figure 3b).

Increased choroid plexus volume has been associated with inflammation and structural alterations by previous studies in AD^85–87^. Although this subtype most often had normal t-tau and p-tau levels (table 1), it had more severe atrophy in comparison to subtype 1, 3 and 5, and had specific atrophy in anterior cingulate areas (figure 3; S-figure 2). Proteins increased in subtype 4 were also enriched for fibroblasts (S-table 5). Fibroblasts produce extracellular matrix proteins, providing structural support to the choroid plexus^84^. Other proteins increased in subtype 4 included cytokines, such as CCL2, CCL21 and CCL15, which can attract monocytes and T lymphocytes ^88^. Of note, proteins with *decreased* levels in subtype 4 were related to axonal outgrowth and synaptic plasticity (e.g., BDNF), in part overlapping with proteins increased in subtype 1 and 2 and also enriched for REST and SUZ12. This suggests that 2 subtype is also characterised by neuronal hypo*-*plasticity.

When testing AD genetic risk variants, we found enrichment in subtype 4 of *ABCA7*, *PICALM,* and *IL34,* and also with *CLNK*. While *ABCA7* and *IL34* are expressed in the choroid plexus ^89, 90^, *PICALM* is expressed in the blood-brain barrier^91^. Both *ABCA7* and *PICALM* play a role in lipid metabolism^92, 93^. Both have been associated with amyloid clearance in combination with LRP1 across the blood-brain barrier (*PICALM*)^94, 95^ or choroid plexus (*ABCA7*), or via lysosomal degradation^92, 96–100^. *IL34* has been associated with impaired macrophage function^101^, which could interfere with macrophage uptake of amyloid fibrils^102^. Of note, this subtype had lower levels of BACE1 and abeta40 than controls (table 1), suggesting *decreased* amyloid metabolism. This suggests that impaired clearance mechanisms underlie subtype 4, rather than amyloid overproduction which was observed in subtype 1 and 2.

Taken together, these results provide further support for choroid plexus dysfunction as another contributor to AD pathogenesis, for a specific subgroup of patients.

### Subtype 5: Blood-brain barrier dysfunction

Subtype 5 (n=56, 13.4%) was highly similar to our previously identified BBB dysfunction subtype with increased levels of 640 proteins that included blood proteins such as albumin, fibrinogens, plasminogen, prothrombin, and many immunoglobulins such as IGG-1, which are all proteins that leak into the brain when the BBB is compromised^103, 104^. Pathways associated with increased proteins included blood coagulation, B cell mediated immunity, and acute inflammatory response. No transcription factor enrichment was observed for proteins with increased CSF levels. On MRI this subtype had more microbleeds than controls (p=.03), unlike the other subtypes (table 1). The majority of proteins associated with subtype 5 (1013, 61%) had, however, *decreased* CSF levels compared to controls, and these were associated with neuroplasticity and converged on transcription factors SUZ12 and REST. This suggests that the BBB subtype, like the choroid plexus subtype, has hypo-plasticity. Neuronal plasticity processes can be impaired by leakage of blood proteins, including fibrin, which were specifically increased in this subtype^104^. Furthermore, in this dataset, we could now detect changes in protein levels that were associated with pericytes, cells that normally cover capillaries. We also found altered levels of proteins associated with particular vascular cell types, such as lower levels of PDGFRB, CDH2 (N-cadherin), MFGE8 (medin), HTRA1, LAMB1 (laminin), EDN1, LRP1, and JAM3, as well as increased levels of CDH5 (VE-cadherin), ANXA3, ICAM1, AMBP, VWF and PTPRB (see S-table 5 for detailed vascular cell annotation). These have been all been associated with deposition of blood proteins in the parenchyma in previous studies^91, 105–109^. The low PDGFRb levels we observed may reflect loss of pericytes, which is in line with brain tissue measures of PDGFRb in rodent models and post-mortem AD^105, 110–112^. Alternatively, the low concentrations we observed in the BBB subtype could reflect loss of other vascular cells such as arterial smooth muscle cells, which normally express PDGFRb^108, 113^. Furthermore, the BBB subtype had, unlike the choroid plexus subtype, *decreased* levels of LRP1, which could indicate reduced pericyte numbers, further impeding amyloid clearance across the BBB^114^.

In terms of genetic risk, this subtype had the highest proportion of *APOE* e4 carriers, albeit not significantly from the other subtypes (table 1). This subtype was further enriched for *IL-34*, *ECHDC3* and *APP. IL-34* risk factor was also associated with the choroid plexus subtype, suggesting it contributes to AD pathogenesis through the vasculature^115^. *ECHDC3* has been associated with lipid metabolism^69^. Some variants in *APP* have been associated with vascular disruption and/or increased occurrence of cerebral amyloid angiopathy (CAA), by producing amyloid fragments that are more difficult to clear^116–118^. The notion that this subtype has BBB dysfunction, suggests that this *APP* variant may contribute to AD risk through vascular integrity.

Together, these data provide new insights into the underlying pathophysiological processes associated with the BBB dysfunction AD subtype.

## Conclusion

In this study we dissected AD disease heterogeneity at a patient level using CSF proteomics, at a level of detail level that approaches the level of complexity achieved in tissue proteomics^3, 6, 8^. Our analyses led to a more in-depth characterisation of three previously identified CSF subtypes (i.e., hyperplasticity, innate immune activation and blood-brain barrier dysfunction), and we identified two new subtypes, one with RNA dysregulation, which showed the most aggressive disease course, and one with choroid plexus dysfunction. Notably, we found that each subtype was associated with distinct AD genetic risk factors, further validating that each CSF AD subtype reflects specific underlying molecular mechanisms. The subtypes also showed profound differences in cortical atrophy patterns, and survival times, underscoring their clinical relevance. Given the distinct patterns of molecular processes and AD genetic risk profiles, it is likely that AD subtypes will require specific treatments. For example, subtype 1 individuals may benefit from TREM2 activating treatments, subtype 2 from innate immune inhibitors, subtype 3 from antisense oligonucleotides that restore RNA processing, subtype 4 from inhibition of monocyte infiltration and subtype 5 from cerebrovascular treatments. At the same time, potential harmful side effects arising from certain treatments may also depend on subtype. For example, while antibodies may more easily cross the blood-brain barrier in subtype 5, these individuals may be at increased risk for cerebral bleeding that can occur with some antibody treatments. Future studies should aim to (re)analyse proteomics in clinical trials to test whether particular treatments may have effects for specific subtypes only. To conclude, CSF based subtyping will be highly useful for the selection of individuals likely to benefit most from a specific therapeutic, either for a priori subject stratification for clinical trials, or for responder and side effect analysis.

## Online Methods

### Participants

This study included individuals from Alzheimer center Amsterdam related studies (i.e., Amsterdam Dementia Cohort (ADC)^16^, EMIF-AD preclin AD^17^ and 90+ studies^18^) and participants who co-enrolled in the ADC biobank and the EPAD study^19^. Participants were selected when they had CSF available and either normal cognition with normal CSF abeta42 as controls (n=187) or if they had abnormal CSF abeta42 (n=419). Among the group with abnormal amyloid levels 107 individuals had intact cognition (i.e., preclinical AD)^119^, 103 had mild cognitive impairment (i.e., prodromal AD)^120^ and 209 had dementia according to international consensus criteria^121–123^. In case more individuals met criteria for inclusion, preference was given to individuals with a known *TREM2* R47H (n=8) or R62H mutation (n=28; see details in *Genetic data* below), and to individuals without dementia who had clinical follow up (n=216).

Information on mortality was available from the Dutch Municipal Register for ADC and EMIF-AD participants. One person for whom proteomics was measured was excluded from further analyses in this study, because this person had normal AD markers in CSF, but a clinical diagnosis of primary progressive aphasia. All studies were approved by local Medical Ethical Committees.

### CSF collection

CSF was collected by lumber puncture between the L3/L4, L4/L5 or L5/S1 intervertebral space with a 25-gauge needle and syringe and collected in polypropylene tubes ^16^. CSF sample processing and biobank storage at the Alzheimer center biobank at the department of Clinical Chemistry was performed according to international guidelines^124^.

### CSF sample preparation for tandem mass tag proteomics

Six hundred and ten samples were stored at −80 °C in seven 96 well plates, 100 µl CSF in each well. Each sample plate was thawed on ice, and 30µg protein (separate 96 well plates containing 40 µl CSF were used to do BCA protein assay on 2×10 µl CSF for protein concentration measurements) from each well was transferred to 1.5 ml protein Low-Bind tubes, and immediately frozen on dry ice. Quality control samples and TMT reference samples were collected from the first plate only since it contained an even mixture of all sample groups. From the first plate, 38 µg of CSF from 93 wells were mixed in a 4 ml glass vial on ice, and 30 µg protein were transferred to 110 1.5ml Protein Low-Bind tubes, 22 QC-samples and 88 TMT reference samples, and immediately frozen on dry ice. All samples were lyophilized using a freeze dryer and kept at −80 °C prior to digestion.

Urea protein digestion were performed as follows. Each day until there were no more samples to process, 28 samples together with one QC sample and two reference samples were added 20 µl 8M Urea/20mM Methylamine, vortexed for 5 min at 1000 rpm and sonicated for 30s in ice cold water. The Urea solution was diluted with 20 µl 50mM TrisHCl/1mM CaCl2 pH 7.6, followed by cysteine reduction (0.4 µmol Dithiotreitol, 1h incubation at RT) and alkylation (1 µmol Iodoacetamide, 1h incubation in the dark at RT). To avoid protease alkylation, excess iodoacetamide were allowed to react with Dithiotreitol by adding 0.08 µmol of the reagent before diluting the Urea to 1M with 50mM TrisHCl/1mM CaCl2 pH 7.6. Trypsin digestions were performed for 16h at 37 °C after adding 0.6mg of the protease (porcine trypsin from Promega, GmbH, Mannheim, Germany).

Sample clean-up was performed using a reverse-phase Oasis 96-well HLB μElution Plate 30 μm (2 mg HLB sorbent, Waters, Milford, MA). After lyophilization, QC samples were resuspended in 25 µl 2% acetonitrile/0.5% formic acid. All other samples were resuspended in 20 µl 50mM HEPES buffer pH 8.2 (4-(2-Hydroxyethyl) piperazin-1-ylethanesulfonic acid) prior to TMT labelling. All samples were vortexed for 30s at 1500 rpm and sonicated for 30s in an ultrasonic bath.

Each reporter in a 5mg TMTpro 16plex reagent set were dissolved in 1ml anhydrous acetonitrile. The 610 samples were labelled in 44 experiments, where each experiment contained 14 samples and 2 reference samples. For each sample/reference sample, 20 µl label was added (the two reference samples were labelled with 126 and 134N in each experiment). The labelling reaction was allowed for 75min before it was stopped by adding 5 µl 5% hydroxylamine. The 16 labelled samples for each experiment were combined and lyophilized (about 240 µg protein), and approximately 150 µg were desalted using a reverse-phase Oasis 96-well HLB Elution Plate 30 μm (10 mg HLB sorbent, Waters, Milford, MA). After lyophilization, the 44 samples were dissolved in 150µl 10mM Ammonium formate pH 7.9, and 65µl were fractionated using an off-line HPLC (Agilent 1260 infinity, Agilent Technologies, Santa Clara, California, USA) equipped with a reversed phase column (XSelect CSH C18, 130Å, 3.5 µm, 1 x 150 mm from Waters Corp, Milford, Massachusetts, USA). Using high pH reversed phase chromatography, peptides were separated during a biphasic Acetonitrile (ACN) gradient from two HPLC pumps (flow rate of 50 µl/min). Solvent A and B were 10mM Ammonium formate pH 7.9 in water and 90% ACN/10% water respectively. The gradient composition was 5%B during trapping (2min) followed by 5-12%B over 1 min, 12–44%B for the next 35min, 44-70%B over 10 min, and 70–95%B over 2min. Elution of very hydrophobic peptides and conditioning of the column were performed for 5 minutes isocratic elution with 95%B and 12 minutes isocratic elution with 5%B respectively.

Peptide were collected in a 500 µl protein Low-Bind 96-well plate during peptide elution, 10 fractions were collected. The first fraction was collected in two wells from 5 to 16 min (5.5 min/well, merged into one fraction), the next 8 fractions (2.7 min/fraction) were collected between 16 and 37.6 min, and the last fraction collected in two wells between 37.6 and 53.6 min (8 min/well, merged into one fraction). Fractions were lyophilized and resuspended in 10 µl 2% ACN/0.5% formic acid (FA), and peptide concentrations were measured on NanoDrop UV-Vis spectrophotometer (Thermo Scientific, Waltham, MA, USA) prior to LC-MS/MS analysis.

### Liquid Chromatography (LC) Tandem Mass Spectrometry (MS) Analysis

About 0.5ug tryptic TMT labelled peptides were injected into an Ultimate 3000 RSLC system (Thermo Fisher Scientific, Waltham, MA, USA) connected online to a Exploris 480 mass spectrometer (Thermo Fisher Scientific, Waltham, MA, USA) equipped with EASY-spray nano-electrospray ion source. Peptides were desalted on a pre-column (Acclaim PepMap 100, 2cm x 75µm ID nanoViper column, packed with 3µm C18 beads) at a flow rate of 5µl/min for 5 min with 0.1% trifluoroacetic acid, before separation on a 50 cm analytical column (PepMap RSLC, 50cm x 75 µm ID EASY-spray column, packed with 2µm C18 beads). During a biphasic ACN gradient from two nanoflow UPLC pumps (solvent A and B were 0.1% FA (vol/vol) in water and 100% ACN respectively), peptides were separated through the reversed phase column at a flow rate of 200 nl/min. The gradient composition was 5%B during trapping (5min) followed by 5-8%B over 1 min, 8–30%B for the next 104min, 30-40%B over 15 min, and 40–80%B over 3min. Elution of very hydrophobic peptides and conditioning of the column was performed for 9 minutes isocratic elution with 80%B and 10 minutes isocratic elution with 5%B. The LC was controlled through Thermo Scientific SII for Xcalibur 1.6.

Peptides eluted from the column were detected in the Exploris 480 Mass Spectrometer (capillary temperature at 275 °C and Ion spray voltage at 2100V) with FAIMS (High field asymmetric waveform ion mobility spectrometry) enabled using two compensation voltages (CVs, −50V and −70V), and “Advanced Peak Determination” on. During each CV, the mass spectrometer was operated in the DDA-mode (data-dependent-acquisition) to automatically switch between one full scan MS and MS/MS acquisition. Instrument control was through Orbitrap Exploris 480 Tune 3.1 and Xcalibur 4.4. The cycle time was maintained at 1.5s/CV. The FAIMS filter performs gas-phase fractionation, enabling preferred accumulation of multiply charged ions to maximize the efficiency of DDA. FAIMS results in less precursor co-isolation, and cleaner MS2 spectra. MS spectra were acquired in the scan range 375-1500 m/z with resolution R = 60 000 at m/z 200, automatic gain control (AGC) target of 3e6 and a maximum injection time (IT) at auto (depending on transient length in the orbitrap). The most intense eluting peptides with charge states 2 to 6 and above an intensity threshold of 2e4 were sequentially isolated to standard target value of 2e5, or a maximum IT of 120 ms in the C-trap, and isolation width maintained at 0.7 m/z (quadrupole isolation), before fragmentation in the HCD (Higher-Energy Collision Dissociation). Fragmentation was performed with a normalized collision energy (NCE) of 32 %, and fragments were detected in the Orbitrap at a resolution of 45 000 at m/z 200, with first mass fixed at m/z 110. One MS/MS spectrum of a precursor mass was allowed before dynamic exclusion for 45s with “exclude isotopes” on. Lock-mass internal calibration was not enabled.

The resulting .raw files were processed using Proteome Discoverer 2.5. The database file used for the search using Sequest HT was Swiss-Prot with 20395 entries (version 20210413.fasta). The following modifications were defined in the database search: Precursor Mass Tolerance: 10 ppm, fragment Mass Tolerance: 0.02 Da, Static Peptide N-Terminus: TMTpro / +304.207 Da (Any N-Terminus), static modification: TMTpro / +304.207 Da (K), static modification: Carbamidomethyl (C), and dynamic modification for Methionine oxidation. Maximum of missed Cleavage Sites was set to 2, with a minimum peptide length of 6. The validation settings were set to 0.01 for target FDR for PSMs and peptides (strict) and 0.05 for relaxed. Peptides used were set to unique + razor. Reporter abundance was based on intensity with a co-isolation threshold of 50 and average reporter S/N threshold of 10. The output results files where then gathered and subjected to further processing.

Technical deviations may influence protein abundances across TMT experiments^125, 126^. We normalized protein abundances according to the Internal Reference Scaling normalization procedure^127^ for TMT proteomic data that uses the common pool reference channels to normalize values between plex experiments, adapted to scale according to median instead of the total sum to reduce sensitivity of outliers. Briefly, this is a two-step approach, with the first step normalizing grand total intensities for each of the 14 channels within an experiment to match these to the two reference channels. In the second step a correction factor is calculated based common pooled internal standards to normalize reporter ion intensities of proteins between TMT experiments. Internal standards were unavailable for 113 proteins, which were excluded for subsequent analyses. S-figure 3 illustrates that the correction effectively removed batch effects.

After batch correction, protein values were log2 transformed, and then scaled according the mean and standard deviation of the control group, such that positive and negative values indicate higher and lower than normal. In total 3863 proteins were identified that had at least 1 observation. For, all proteins we report GENE names to aid comparisons with other AD subtyping literature using either proteomics or RNAseq data.

### CSF ELISA measures

Abeta42, t-tau and p-tau were previously determined using ELISA assays from Innotest (Fujirebio, formerly Innogenetics) in the ADC^128, 129^, or with the Roche Elecsys System (n=15 from ADC and in EPAD^130^. In EMIF-AD preclinAD study amyloid status was determined based on the abeta42 and abeta40, t-tau and p-tau181 were measured with ELISAs from ADx Neurosciences/EUROIMMUN^131^. In EMIF-AD 90+ amyloid status was determined with visual read of [^18^F] flutemetamol positron emission tomography^132^. For these individuals (n=22) tau levels were computed from the TMT MAPT measures, which correlated strongly (r=0.81, p<.001) with Innotest t-tau levels in the ADC cohort (formula: Inno t-tau = - 309.16+0.01*MAPT). For tau categorization, we used t-tau values as these were available in all individuals, and correlated strongly with p-tau levels (r=0.94, p<.001). We used published cutoffs to label individuals as having a normal AD CSF profile or abnormal amyloid based on CSF^128–131, 133^. Three individuals with normal cognition had normal amyloid and abnormal CSF t-tau levels, which were excluded from the present analyses, resulting in a final sample size of 187 controls and 419 individuals with abnormal amyloid. We standardized continuous amyloid 1-42, t-tau and p-tau181 values within specific assays according to the mean and standard deviation of controls to compare these values between subtypes. Finally, we measured BACE1, abeta40, and neurogranin with EUROIMMUN ELISA assays (Germany), NEFL with ADx NeuroSciences (Belgium) ELISA assay, and VAMP2 with a prototype assay developed by ADx Neurosciences (Belgium) with single-molecule array (Simoa) technology (Quanterix Corp, Billerica, USA) as previously described^134^. These measures were not included in clustering, but used as independent markers to compare between AD subtypes.

### Genetic data

*APOE* genotyping was performed in blood as previously described^16, 17, 135, 136^. A subset of 560 individuals we had genotyping data available (Illumina Global Screening Array, GSA). Details on quality control procedures were previously described^137^, and for EPAD available on GitHub (https://github.com/marioni-group/epad-gwas). Genotype vcf files were imputed using the TopMed reference panel^138^. Eighty-three genetic risk loci for AD were selected based on their previous genome-wide association with AD^5^. These SNPs were extracted from the genetic data based on rsID/and or base pair location in the genome. Protective SNPs (i.e., odds ratios below 1) were inverted, such that higher values indicate more AD risk for all SNPs.

### MRI data

A subset 503 individuals had structural T1 weighted MRI available. To test if subtypes were characterized by different atrophy patterns, we restricted analyses to subtypes in the dementia stage (n=159 and 160 controls), because in that stage atrophy is most pronounced. Acquisition details were previously described^17, 132, 139, 140^. Cortical thickness, hippocampal volume, choroid plexus volume and total intracranial volumes were estimated with FreeSurfer version 7.1.1 (http://surfer.nmr.mgh.harvard.edu/)^141^. Cortical thickness and volumetric estimates were summarised in anatomical regions as defined by the FreeSurfer implementation of the Desikan-Killiany atlas. Choroid plexus and hippocampal volumes were adjusted for total intracranial volume to adjust interindividual differences in head size. Furthermore, microbleeds were counted on T2* sequences by an experienced neuroradiologist and defined as small round hypointense foci up to 10mm in the brain parenchyma^17, 132, 139, 140^.

### AD subtype discovery

Our objective was to identify subtypes *within* AD, and so we first selected proteins that were related to AD. For this, we compared all AD individuals to controls on CSF levels of proteins that were observed in the complete sample with Kruskal-Wallis tests. Because previous studies have indicated that AD related alterations of CSF protein levels may depend on cognitive states and/or tau status in a non-linear way^9, 13, 142^, we repeated these analyses stratified for these factors. This resulted in 1058 proteins that were selected for clustering with non-negative matrix factorization as implemented in the ‘NMF’ package^143^ v0.25 in R version 4.2.2. “Bird Hippie”. We followed the procedure as in our previous study^9^. Briefly, proteins were first scaled to have positive values between 1 and 2, keeping relative values intact. Next, we performed 30 different runs of NMF to determine the number of clusters (i.e., subtypes) that best described the data. We tested up to 10 clusters, and found that 5 clusters showed an optimal balance of a high co-phonetic coefficient, at least 2-fold improved fit over lower clustering solution as compared to improvement by random cluster solution, and a silhouette score of >0.5 (supplemental table 3). We then labelled each individual patient according to the subtype that best matched their proteomic profile. Patient level subtype clusters were visualized by projecting the NMF subtype scores to a UMAP embedding, via construction of a k-nearest neighbor graph using the ‘uwot’ R package (v 0.1.14). Patient level subtype labels provide the basis for all subsequent post-hoc comparisons, as described in the next sections.

### Biological characterization of AD subtypes

We characterized the biological processes associated with AD subtypes, by comparing the subtypes on CSF protein levels of all available proteins, including in addition to the fully observed proteins also proteins with missing values when they had at least 5 observations available in each subtype group (2907 proteins in total). For this we used linear models with participant subtype status as predictors and protein levels as outcomes. We repeated analyses correcting for age and sex, and stratifying according to cognitive state to determine influence of these factors on the results. All subtypes were compared to the control group, as well as to each other, and results from all comparisons are reported in supplementary table 5. In the main manuscript we report results of comparisons to controls only. To test the biological pathways associated with subtypes, we performed pathway enrichment analyses for biological processes from the Gene Ontology (GO) release 2022-01-13 as accessed by Panther^144^ version 16.0, for the proteins that were associated with each subtype (i.e., differed from controls with p<.05), separately for increased and decreased alterations. Hypergeometric Fisher’s exact test were used for pathway enrichment, and pathway p values were corrected for multiple testing with the false discovery rate procedure. We further tested if AD subtype related proteins were associated with potential up stream transcription factors from the CHEA and ENCODE databases through ENRICHR^145^. We further annotated proteins for cell type specificity according to the Human Protein Atlas (https://www.proteinatlas.org), and the RNAseq Barres database^146^; for specific vascular cell types with Garcia et al., (2022)^108^ and Yang et al., (2022)^91^; for choroid plexus associations according to the Harmonizome database^147^; for REST signalling associations based on Meyer et al., (2019) ^35^ and Otero-Garcia et al., (2022)^148^; for blood-brain barrier dysfunction according to Dayon et al., (2019)^103^; and for CSF pathway panels informed by on tissue proteomics to Higginbotham et al, (2020)^4^.

### Post hoc comparisons between subtypes on clinical, MRI and genetic characteristics

We performed post-hoc tests to characterize AD subtypes clinically and biologically with Chi^2^ tests for discrete variables (sex and APOE e4 genotype), and with linear regression models for continuous variables correcting for age and sex when applicable. Subtype differences in time to progress to dementia was tested with Cox proportional hazard models, and restricted to individuals the prodromal stage for reasons of statistical power. Subtype differences in survival times were also tested with Cox proportional hazard models, and restricted to individuals in the dementia stage for reasons of statistical power. Subtype differences with controls on genetic variants (i.e., SNPs, as continuous outcome) were tested with linear regression models, taking imputation uncertainty into account when possible, and repeated including age and sex as covariates. Subtype differences in hippocampal volume was tested in all stages, because this structure is altered in very early clinical stages.

## Data Availability

All data produced in the present study are available upon reasonable request to the authors

## Acknowledgements

Acknowledgements/funding

BMT: ZonMW VIDI #09150171910068; the Dutch L’Óreal-UNESCO fellowship 2022 for women in science;

PJV: ZonMW Redefining AD #733050824 received support from the EU/EFPIA Innovative Medicines Initiative Joint Undertaking (EPAD grant n° 115736; EPND grant n°101034344), and from the Innovative Medicines Initiative 2 Joint Undertaking (JU) under grant agreement number 101034344 (EPND). The IMI JU receives support from the European Union’s Horizon 2020 research and innovation programme and EFPIA.

LV: received research support from ZonMW, Alzheimer Nederland and Stichting Dioraphte. WF is supported by the Pasman stichting. WF is recipient of OTAPA, a collaboration project which is co-funded by the PPP Allowance made available by health-Holland, Top Sector Life Sciences & Health to stimulate public-private partnerships and Brain Research Center (grant no. LSHM19051). WF is recipient of JPND-funded E-DADS (ZonMW project # 733051106). WF is recipient of ABOARD, which is a public-private partnership receiving funding from ZonMW (#73305095007) and Health∼Holland, Topsector Life Sciences & Health (PPP-allowance; #LSHM20106). VV is supported by JPND-funded E-DADS project (ZonMW project #733051106).

LMR was funded by a ZonMW Memorabel fellowship (#10510022110012).

SL is recipient of ZonMW funding (#733050512). Genotyping of the Dutch case-control samples was performed in the context of EADB (European Alzheimer DNA biobank) funded by the JPco-fuND FP-829-029 (ZonMW projectnumber 733051061.

CET is supported by the European Commission (Marie Curie International Training Network, grant agreement No 860197 (MIRIADE), Innovative Medicines Initiatives 3TR (Horizon 2020, grant no 831434) EPND (IMI 2 Joint Undertaking (JU), grant No. 101034344) and JPND (bPRIDE), National MS Society (Progressive MS alliance), Alzheimer Association, Health

Holland, the Dutch Research Council (ZonMW), Alzheimer Drug Discovery Foundation, The Selfridges Group Foundation, Alzheimer Netherlands. CT is recipient of ABOARD, which is a public-private partnership receiving funding from ZonMW (#73305095007) and Health∼Holland, Topsector Life Sciences & Health (PPP-allowance; #LSHM20106).

Research of the Alzheimer center Amsterdam is part of the neurodegeneration research program of Amsterdam Neuroscience. Alzheimer Center Amsterdam is supported by Stichting Alzheimer Nederland and Stichting VUmc fonds. For part of this work the Dutch national e-infrastructure was used with the support of the SURF Cooperative using grant no. EINF-2044.

Mass spectrometry-based proteomic analyses were performed by FBerven at the Proteomics Unit at the University of Bergen (PROBE). This facility is a member of the National Network of Advanced Proteomics Infrastructure (NAPI), which is funded by the Research Council of Norway (INFRASTRUKTUR-program project number: 295910).

## Competing interests

PJV and BMT co-inventors on patent of CSF proteomic subtypes (published under US2022196683A1, owner Stichting VUmc).

EVM is co-founder of ADx NeuroSciences, while JG is an employee of ADx NeuroSciences.

FB reports editorial fees from Springer, consulting fees from Biogen, IXICO Ltd and Combinostics, steering committee and DSMB compensation from Prothena, USC-ATRI, Merck, Biogen and grants from Roche, Merck, Biogen, IMI-EU, GE Healthcare, UK MS Society.

LV received consulting fees from Roch and Olink, all paid to Amsterdam UMC.

CET has a collaboration contract with ADx Neurosciences, Quanterix and Eli Lilly, performed contract research or received grants from AC-Immune, Axon Neurosciences, BioConnect, Bioorchestra, Brainstorm Therapeutics, Celgene, EIP Pharma, Eisai, Fujirebio, Grifols, Instant Nano Biosensors, Merck, Novo Nordisk, PeopleBio, Roche, Siemens, Toyama, Vivoryon.

She is editor of Alzheimer Research and Therapy, and serves on editorial boards of Medidact Neurologie/Springer, and Neurology: Neuroimmunology & Neuroinflammation.

