## Supplementary material for "Large-scale cerebrospinal fluid proteomic analysis in Alzheimer’s disease patients reveals five molecular subtypes with distinct genetic risk profiles": S-figure 1

Supplementary figure 1

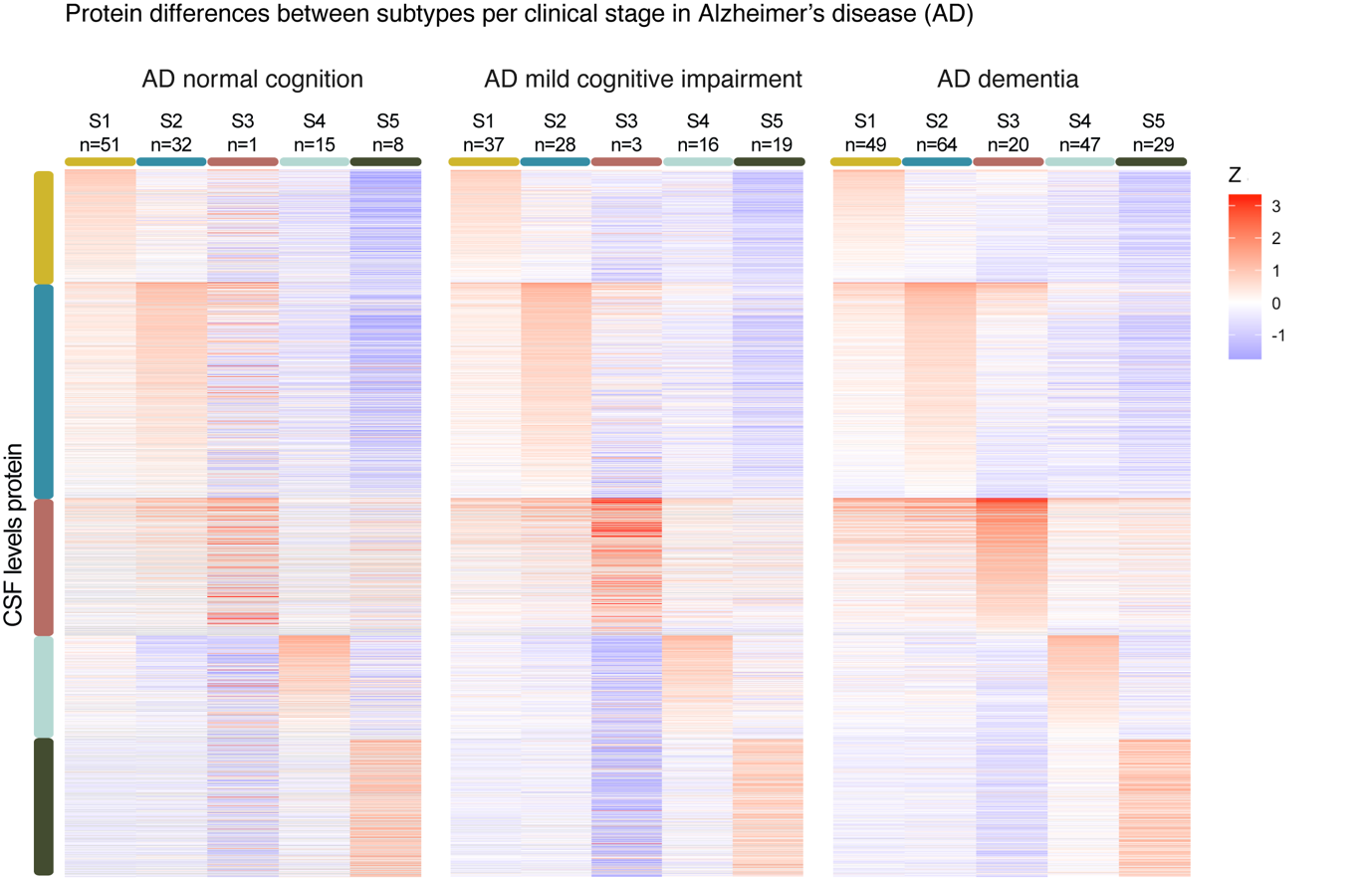

**Supplementary figure 1.** Comparing Alzheimer’s disease (AD) subtypes on protein levels against controls, plotted separately according to clinical stage indicates highly similar subtype patterns, suggesting that protein levels reflect particular AD related traits. See supplementary table 5 columns CY to HU for statistical metrics of subtype comparisons within each clinical stage. All proteins were scaled according to the mean and standard deviation of the control group, such that positive values indicate higher levels than controls, and negative values lower levels than controls.

Supplementary figure 2

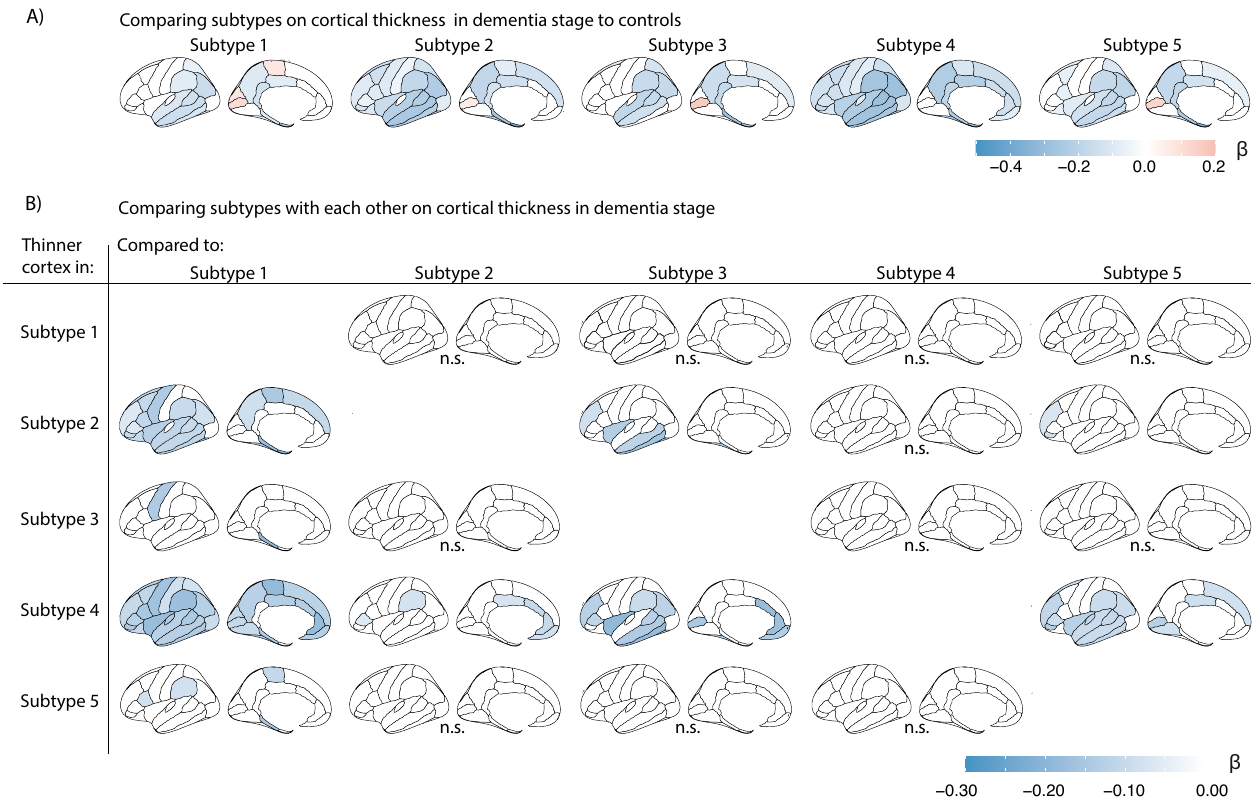

**Supplementary figure 2.** Comparing subtypes on cortical thickness in dementia stage **A)** Cortical thickness compared between AD subtypes (in dementia stage) with controls. **B)** Cortical thickness comparisons between AD subtypes within the dementia stage. Negative values indicate thinner cortex in the subtype indicated in the row as compared to the subtype indicated in the column. Analyses were adjusted for age and sex.

Supplementary figure 3

a)

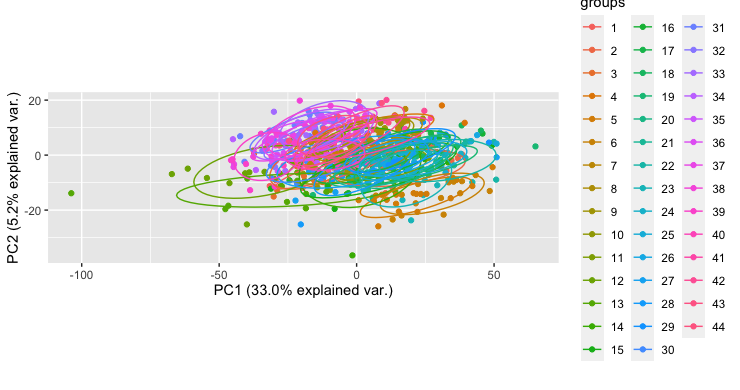

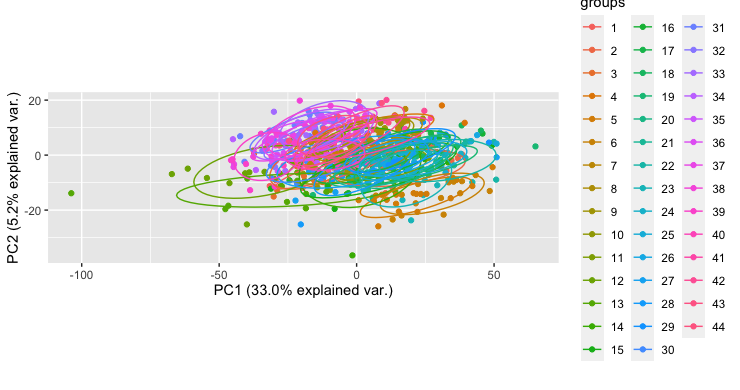

b)

TMT experiment

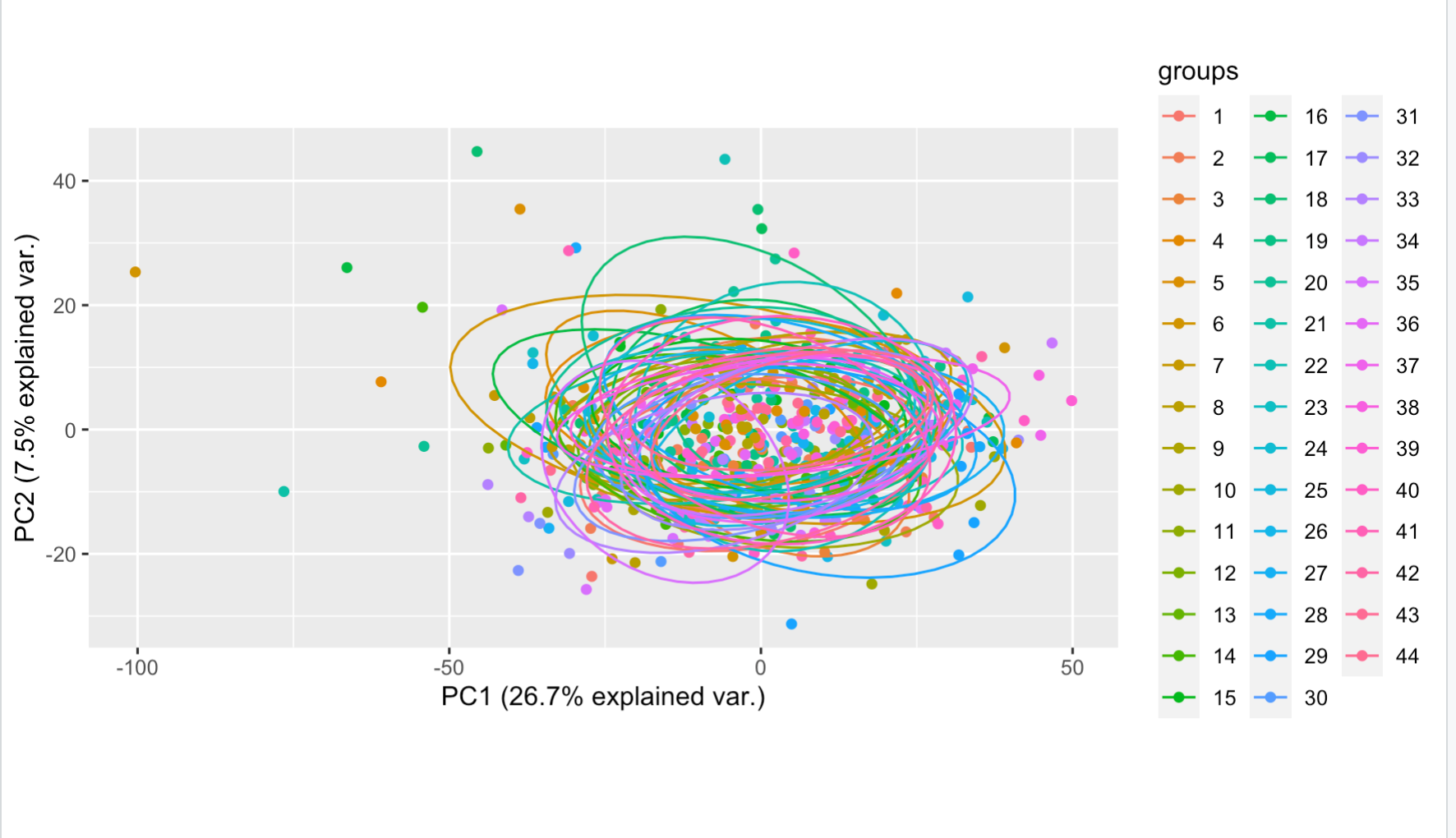

**Supplementary figure 3** Batch correction of TMT experiments. a) Biplot of first two principal components on unnormalized protein abundances have batch effects between TMT experiments as indicated by the non-overlapping circles that correspond to all 44 TMT experiments. b) Batch effects were successfully removed with the Internal Reference Scaling method as described in the online methods.
